# Don’t put words in my mouth: Speech perception can generate False Positive activation of a speech BCI

**DOI:** 10.1101/2024.01.21.23300437

**Authors:** A. Schippers, M.J. Vansteensel, Z.V. Freudenburg, N.F. Ramsey

## Abstract

Recent studies have demonstrated that speech can be decoded from brain activity and used for brain-computer interface (BCI)-based communication. It is however also known that the area often used as a signal source for speech decoding BCIs, the sensorimotor cortex (SMC), is also engaged when people perceive speech, thus making speech perception a potential source of false positive activation of the BCI. The current study investigated if and how speech perception may interfere with reliable speech BCI control. We recorded high-density electrocorticography (HD-ECoG) data from five subjects while they performed a speech perception and speech production task and trained a support-vector machine (SVM) on the produced speech data. Our results show that decoders that are highly reliable at detecting self-produced speech from brain signals also generate false positives during the perception of speech. We conclude that speech perception interferes with reliable BCI control, and that efforts to limit the occurrence of false positives during daily-life BCI use should be implemented in BCI design to increase the likelihood of successful adaptation by end users.

## Introduction

Neurodegenerative conditions such as amyotrophic lateral sclerosis (ALS) can lead to severe paralysis, to the point where individuals enter a locked-in state; they are completely unable to voluntarily move and speak but remain cognitively intact (American Congress of Rehabilitation Medicine, 1995; Hayashi & Kato, 1989). For these individuals, a Brain-Computer Interface (BCI) may provide a new means of communication. A BCI allows a user to control a computer by modulating their brain activity, for example by attempting to move their hand or to produce speech. In this way, an individual with severe paralysis can operate a device and spell letters, words, or even produce full sentences.

Recent developments in the field of implanted BCIs have demonstrated their huge potential as a communication solution for people with severe motor impairment. It has been demonstrated that BCIs can be used by individuals with ALS to control a communication application on a computer by means of a binary click based on attempted hand movements (Oxley et al., 2021; Vansteensel et al., 2016), with long-term stability (Pels et al., 2019). Another way to control a device is through independent multidimensional cursor navigation based on the production of speech commands (Luo et al., 2023). Others have focused on direct speech decoding for communication. Using subdural electrocorticography (ECoG) grids to record cortical activity during (attempted) speech production, it was shown that it is feasible to decode words and full sentences in a limited vocabulary (Moses et al., 2021) and to decode sentences in a large vocabulary by spelling individual letters (Metzger et al., 2022). ECoG-based activity patterns during speech production can also directly be synthesized into speech sounds (Angrick et al., 2023; Herff et al., 2019), or combined with the decoding of discrete words into a multimodal speech prosthesis (Metzger et al., 2023). More invasive approaches such as microelectrode arrays can also be used to control a speech BCI, by decoding speech in a large vocabulary with high accuracy and a high rate of decoded words per minute (Willett et al., 2023), or by directly synthesizing speech (Wairagkar et al., 2023).

Incorporating the needs and wishes of potential end-users is of utmost importance for successful adoption of BCIs in the daily lives of these users. Efficiency and user satisfaction are two factors that affect usability, but especially in situations where a BCI is used for communication and device control, high effectiveness of the device is crucial (Kübler et al., 2014). Accuracy and communication speed are two areas that are heavily investigated as demonstrated by the studies described above, but reliability in real-life settings has received too little interest. A reliable BCI is one that decodes each communication-attempt that is intended (true positives) and remains silent when the user is not intending to communicate (true negatives). A BCI that is unreliable would miss communication attempts (false negatives) and produces BCI activations or utterances when the user was not intending to do so (false positives). In the case of speech BCIs, false positives may occur when the brain activity patterns outside of attempts to speak resemble those during speech production more than they resemble a pattern of baseline rest activity. If a BCI is too sensitive and also labels non-intended brain activity changes as communication attempts, the resulting false positives may be a source of annoyance by both the user and their caregivers since the respective BCI utterances can interrupt normal conversation or cause confusion about the needs and wishes of the user. A high false positive rate may even cause caregivers to ignore BCI output if they experience that a large number of utterances are in fact not attempted by the user. Designing an accurate system that retains a low false positive rate in all circumstances is thus of utmost importance for speech BCIs to be successfully adopted in daily life.

Most speech BCIs are based on brain signals originating from the sensorimotor cortex (SMC, pre- and postcentral gyri), of which the most ventral part is activated during the movement of the articulators (Bouchard et al., 2013). However, this area is known to also become activated during the perception of speech (Cheung et al., 2016; Pulvermüller et al., 2006; Rhone et al., 2016; Skipper et al., 2005; Venezia et al., 2021). Since speech BCIs utilize the cortical activation patterns originating from the SMC to decode produced or attempted speech, and since this area also activates during the perception of speech, speech perception is a potential source of false positive activation of a speech BCI.

Considering speech perception is a vital part of human interaction and communication, it is important to verify if and how it may interfere with reliable speech BCI control. Therefore, the current study investigates the similarities between brain activity patterns during speech perception and production, and tests whether decoders designed to detect and classify produced speech generate false positives during the perception of speech. For this, commonly used signal features for speech decoding were extracted from high-density (HD) ECoG recordings stemming from the SMC of five participants. Given that end-users of BCI implants for communication will not be able to generate sounds, we conducted the analyses for overt as well as for mimed speech. First, we identified which areas of the SMC are responsive to speech perception and/or speech production. Second, we trained and tested a decoder on the speech production data to determine decoding performance, and then tested the same decoder on the speech perception data to investigate the false positive activation of the decoder by speech perception. To account for fluctuations in brain signal amplitude across days, which may necessitate daily calibration of a BCI, we conducted the analyses using two normalization methods (based on an independent rest period and on rest data acquired during BCI experiments). Finally, we investigated the differences between the brain activity patterns during speech perception and production, which could pave the way for a strategy to limit the occurrence of false positives in daily-life speech BCI use.

## Methods

### Participants

Five participants (mean age 33.4 years, two females, all right-handed, see Table 1) who underwent epilepsy treatment at the University Medical Center Utrecht were included in the current study. As part of their presurgical assessment, they were implanted with subdural clinical ECoG electrode grids. They gave written informed consent to participate in this study and for the implantation of an additional HD electrode grid, which was placed over the SMC. Only the ECoG data acquired with these HD grids are used in this study. The study was conducted in accordance with the Declaration of Helsinki (2013) and approved by the Medical Ethical Committee of the University Medical Center Utrecht.

**Table 1:** Participant demographics and timeline of data acquisition.

Day 0 is day of electrode grid implantation.
|  | Age range at implant | Sex | Speech perception recording | Speech production recording | Rest recording |
| --- | --- | --- | --- | --- | --- |
| 1 | 21-25 | M | Day 2 | Day 2 | Day 3 |
| 2 | 36-40 | F | Day 4 | Day 7 | Day 5 |
| 3 | 30-35 | M | Day1 | Day 1 | Day 2 |
| 4 | 46-50 | F | Day 2 | Day 3 | Day 3 |
| 5 | 26-30 | M | Day 1 | Day 2 | Day 2 |

### Task design

All participants performed a speech perception and a speech production task, which both included the same sequence of speech utterances: “Do”, “Re”, “Mi”, “Fa”, “So”, “La”, and “Ti”. This sequence of speech sounds was chosen since it was (expected to be) intrinsically known to the participants (thus not needing stimulus presentation), therefore limiting the cognitive processes occurring during the tasks. One trial comprised the full sequence of seven sounds. Both speech tasks were about 9 minutes in duration.

In the speech perception task, the speech stimuli were presented in an audiovisual, visual-only, and audio-only fashion. In the audiovisual trials, the lower half of a women’s face was presented on the screen while she produced the speech utterances, with about 1.75 seconds between utterances. In the visual-only condition the same video was shown but there was no sound, while in the audio-only condition a still of the woman’s face in resting position (mouth closed) was presented together with the audio. Each perception condition was presented ten times, in random order, and perception trials were alternated with rest trials, indicated by a fixation cross which lasted 2.5 seconds. Participants were instructed to attentively watch the screen and listen to the sounds, but not produce any movements or sounds themselves. For this study, only the audiovisual perception trials were used in addition to the rest trials, since these best simulate a real-life communication setting. The rest trials corresponding to this task will further be referred to as the perception-rest trials.

In the speech production task, participants were instructed to produce the same sequence of utterances (each complete sequence constituting one trial). The task was presented on a laptop placed about 1m in front of the participant. Prior to each sequence, an instruction on the screen (presented for 1.5 seconds) indicated if participants should produce the sequence in an overt, whispered, or mimed manner. Each condition was presented ten times, in random order. Participants were cued to produce the sequence of sounds with 1.75 seconds intervals, using a rotating cursor. The first speech utterance (“Do”) was cued 1.75 seconds after the instruction. Participants were instructed to produce the sounds monotonally. Speech production trials were alternated with rest trials, which were indicated by a fixation cross and lasted 2.5 seconds. In this study, only the overt and mimed production trials were used, in addition to rest, since these manners of speech production are used most in speech decoding or emulate the situation of a locked-in end-user. The rest trials corresponding to this task will further be referred to as the production-rest trials.

A rest task was recorded in addition to the speech tasks. During this rest task, which lasted three minutes, participants were instructed to focus on a fixation cross presented on a screen placed about one meter in front of them while refraining from generating movements.

### Electrode locations

The HD ECoG grids were placed over the face area of the left SMC, which was close to, but outside of the area of clinical interest. After implantation, a pre-implantation MRI and a post-implantation CT scan were used to determine the exact location of the grids (using the ALICE toolbox, Branco et al., 2018). Using Freesurfer surface reconstructions of each individual brain and the electrode coordinates, each electrode was assigned to an anatomical region according to the Allen atlas (see Figure 1 for electrode placements and Table 2 for information on the implanted grids). Electrode coordinates were converted to Montreal Neurological Institute (MNI) space to allow for anatomical comparison between participants.

**Figure 1:**
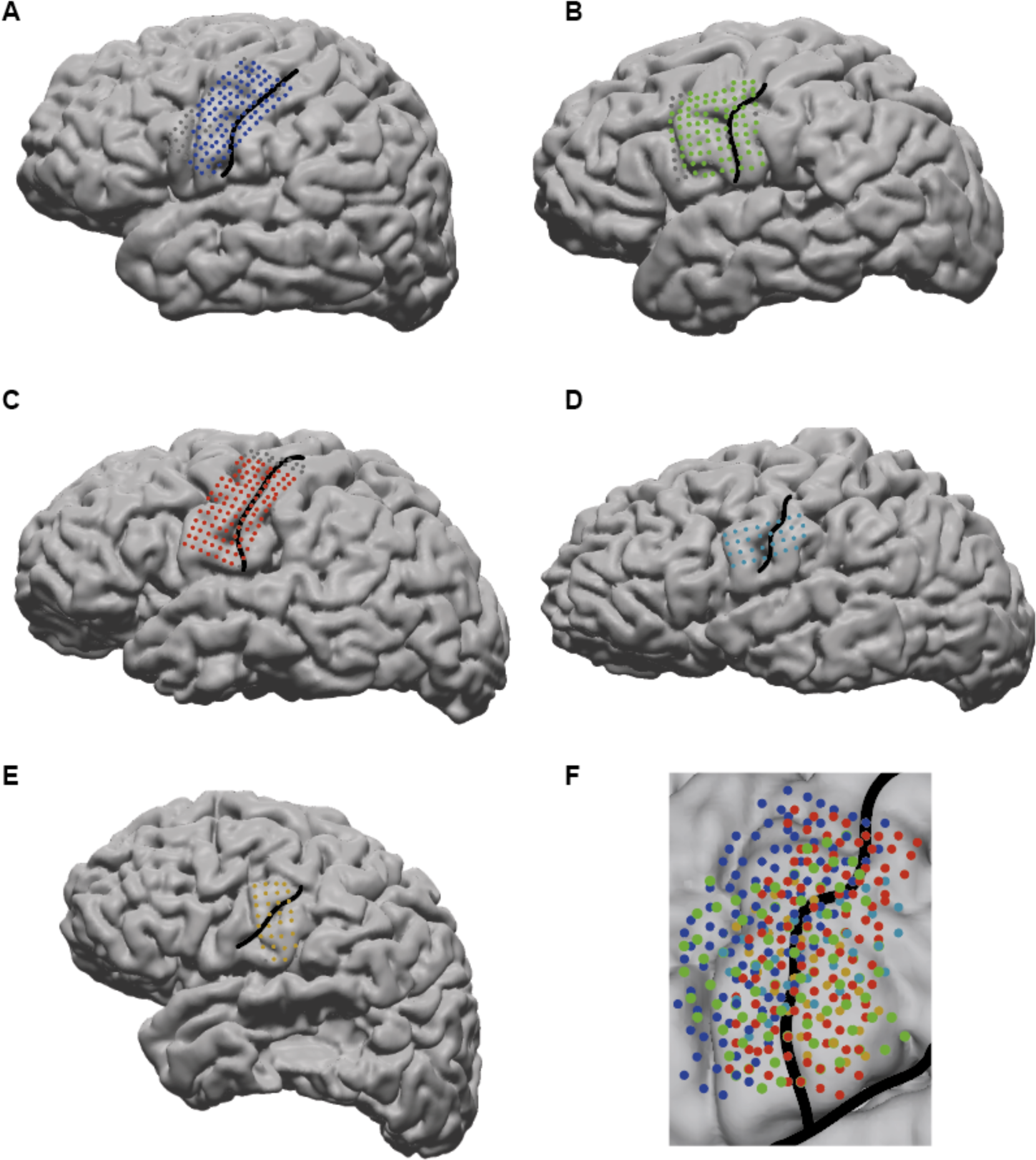
Grid placement. A-E) Grid placement on individual surface reconstructions for participants 1 – 5. Central sulcus is indicated with a black line. F) Electrode placement (coordinates converted to MNI space) of all included electrodes of all participants, projected to an MNI cortex reconstruction. Central sulcus and Sylvian fissure are indicated with black lines.

**Table 2:** Grid and electrode information.

|  | Grid manufacturer | Exposed electrode diameter | Inter-electrode distance | # Total electrodes | # Excluded electrodes due to poor signal quality | Final # included SMC electrodes |
| --- | --- | --- | --- | --- | --- | --- |
| 1 | PMT Corporation | 1 mm | 3 mm | 128 | 9 | 105 |
| 2 | AD-Tech Medical Instrument Corporation | 1.17 mm | 3 mm | 96 | 3 | 84 |
| 3 | PMT Corporation | 1 mm | 3 mm | 128 | 18 | 108 |
| 4 | CorTec | 1 mm | 4 mm | 32 | 0 | 31 |
| 5 | CorTec | 1 mm | 4 mm | 32 | 1 | 30 |

### Data acquisition

For participants 1 – 3, a 256-channel Blackrock Neuroport system (Salt Lake City, USA) was used to record the HD-ECoG signals at a sampling frequency of 2000 Hz (band pass filtered between 0.3 – 500 Hz). Audio was recorded simultaneously on the Neuroport system, at a sampling frequency of 30 kHz. For participants 4 and 5, the HD-ECoG was recorded on a Micromed system (Treviso, Italy) at a sampling frequency of 2048 Hz (band pass filtered between 0.15 – 926.7 Hz), and audio was recorded in the task presentation software (Presentation Neurobehavioral Systems) at 44.1 kHz. For all participants, the brain recordings were referenced to a clinical reference and a ground electrode placed on the forehead and mastoid. Video recordings were made on the Micromed system during task execution and video, audio, and HD-ECoG recordings were aligned using a beep presented at the start of each task and an event marker that was sent to the recording system.

### Data preprocessing and feature extraction

The HD-ECoG data was tested for acoustic contamination following the methods provided by (Roussel et al., 2020). No signs of acoustic contamination were found in the speech perception and production data for all participants.

The data was preprocessed by first applying a notch filter to remove line noise at 50 and 100 Hz. Second, electrodes that were located on top of a clinical grid, facing the dura, or those showing poor signal quality (e.g., excessive noise, flat signals) were identified and excluded from further analysis (see Table 2 for number of electrodes per participant). Third, a common average re-reference (CAR) was applied on the remaining electrode signals. The resulting preprocessed data was downsampled to 500 Hz.

To extract the spectral response for each electrode, a Gabor Wavelet Dictionary was used. The ECoG signal was convolved with Gabor wavelets with a full-width half-maximum of four wavelengths in each individual frequency from 1 to 130 Hz. To compute the power response within the high-frequency band (HFB) the log of the sum of the absolute values was taken within the frequency range of 65 to 95 Hz. The resulting power responses were normalized by z-scoring the signals using the mean and standard deviation from all included electrodes over the three-minute rest task. Finally, the power responses were smoothed over a window of 50 ms. Only the electrodes showing good signal quality in all tasks (speech perception, speech production, and rest) and that were positioned over the SMC (pre- and postcentral gyrus) were included in further analysis.

### Speech epoch extraction

For each individual speech stimulus, the power trace within a period of interest around speech onset was extracted from the HFB power signals, which will be referred to as a speech epoch. For the perception data, stimulus onset was defined as the moment the speech sound could be audibly perceived. The period of interest for the speech perception task is 0 to 700 ms after stimulus onset. For the speech production task, speech onset was defined as the first moment the speech sound could audibly be perceived. For the mimed sounds, the speech onset was determined as the first moment a speech movement could be observed on the video recordings. The period of interest for the speech production task was 200 ms before until 500 ms after speech onset.

A maximum of 70 active speech epochs were extracted from the power data for each task. Production epochs in which participants did not produce the correct sound were excluded (six and seven overt epochs for participants 2 and 4, respectively, five, eleven and one mimed epochs for participants 1, 2, and 4, respectively). No speech perception epochs had to be excluded.

In the speech production and perception tasks, seven rest epochs of 700 ms were extracted from each 2.5 second rest period by randomly sampling seven time periods that fell within the 2.5 second rest period. Doing this, a maximum of 210 rest epochs could be extracted per task (less if there was noise or movement during rest epochs: seven perception-rest epochs for participants 1, 4, and 5; seven production-rest epochs for participants 4 and 5, 42 production-rest epochs for participant 2). The resulting imbalance of epoch numbers between active (perception/production) and rest epochs was maintained since during daily-life use, we expect a similar disbalance in moments where a user is intending to communicate speech versus when they are not intending to communicate speech.

### Speech-related cortical engagement

To determine if there were speech-related activity changes in the SMC during each speech task, the power signals during active trials (perceived or produced speech) were compared to the power signals during the rest periods of that task. An R^2^ was computed for each electrode and each task condition separately by correlating the mean response over time in each speech trial to the task condition (zeros for the rest trials versus ones for the active trials). For the active trials, this was the mean power over all seven epochs of 700 ms within that trial, for the rest trials this was the mean over the entire 2.5 second rest periods. The effect size of the (de)activation of the power response was expressed as a signed R^2^, where a high positive R^2^ indicates a strong power increase during active periods as compared to rest. The significance of the R^2^ values was determined at p < .05, Bonferroni corrected for the number of included SMC electrodes.

### Classification of produced and perceived speech epochs

To test whether a speech production-based decoding algorithm would generate false positives during speech perception, first a support vector machine (SVM) decoder needed to be trained that could accurately decode produced speech sounds. This was done separately for the overt and the mimed produced epochs. For this purpose, the mean power response over time per electrode was calculated for each production and production-rest epoch. Then, a Leave-one-Out (LoO) approach was used to decode each epoch (eight-way classification: seven speech sounds and rest). Specifically, an SVM was trained on all but one epoch, and then tested on the left-out epoch. After doing this for all individual production (active and rest) epochs, decoding accuracy could be calculated by dividing the number of correctly classified epochs by the total number of epochs and multiplying that by 100 to get a percentage score. To determine if classification accuracy was significantly above chance level the production labels were randomly shuffled, and production epochs were again classified using those shuffled labels with a LoO approach. This was repeated a total of 1000 times, which generated a distribution of random classification scores. The classification accuracy using the unshuffled labels was compared to the random distribution and considered significantly above chance if it fell in the upper 5% (p < .05) of random classification scores.

To test whether a decoder based on produced speech would generate false positives during perception, an SVM was trained on all production (active and rest) epochs, again separately for the overt and the mimed produced epochs. All speech perception and perception-rest epochs were classified based on this decoder. All perception and perception-rest epochs that were classified as anything else than a rest epoch were considered false positives. The percentage of false positives was calculated by dividing the number of non-rest classifications by the total number of epochs and multiplying this by 100.

To verify whether the false positive epochs were caused by baseline differences in the task recordings (caused by for example day-to-day variations in power amplitudes), all epochs were normalized based on the rest data corresponding to each task. For this, the mean and standard deviation was calculated per electrode over all 2.5 second rest periods within each speech task. Then, each epoch was z-scored by subtracting the mean from each timepoint and dividing this by the standard deviation. This rest normalization brought the power signals from different tasks within the same range. The same steps as described above were then taken to determine the classification accuracy on production epochs (and significance of this score) and the percentage of false positives during the perception epochs, now using the z-scored epochs.

### Distinguishing speech perception from speech production and rest

To investigate the differences between brain activity patterns during speech perception and speech production, we tested if the two brain states could be distinguished from each other and from rest. This was done on the rest-normalized epochs, since after rest normalization the power amplitudes from the different task recordings were brought to the same range, allowing the rest epochs from the two different tasks to be grouped. Using a LoO approach, each individual epoch was classified as being a speech perception, speech production or rest epoch, based on an SVM trained on all other epochs. Classification accuracy was determined by dividing the number of correctly classified epochs by the total number of epochs and multiplying this by 100 to get a percentage score.

## Results

### Speech-related cortical engagement

All participants showed engagement of the SMC during both speech perception and production, as quantified by having at least one electrode with a significant increase in HFB power during the active periods compared to rest. The responsive areas partially overlapped between the two tasks (see Figure 2). Of the electrodes that had a significant increase in HFB power during speech perception, on average 67.14% were also significantly activated during overt speech production (3/3, 2/4, 6/7, 0/1, and 2/2 electrodes for participants 1 – 5, respectively). In addition, an average of 65.71% of electrodes that were significantly activated during speech perception were also activated during mimed speech production (3/3, 0/4, 2/7, 1/1, and 2/2 electrodes for participants 1 - 5, respectively).

**Figure 2:**
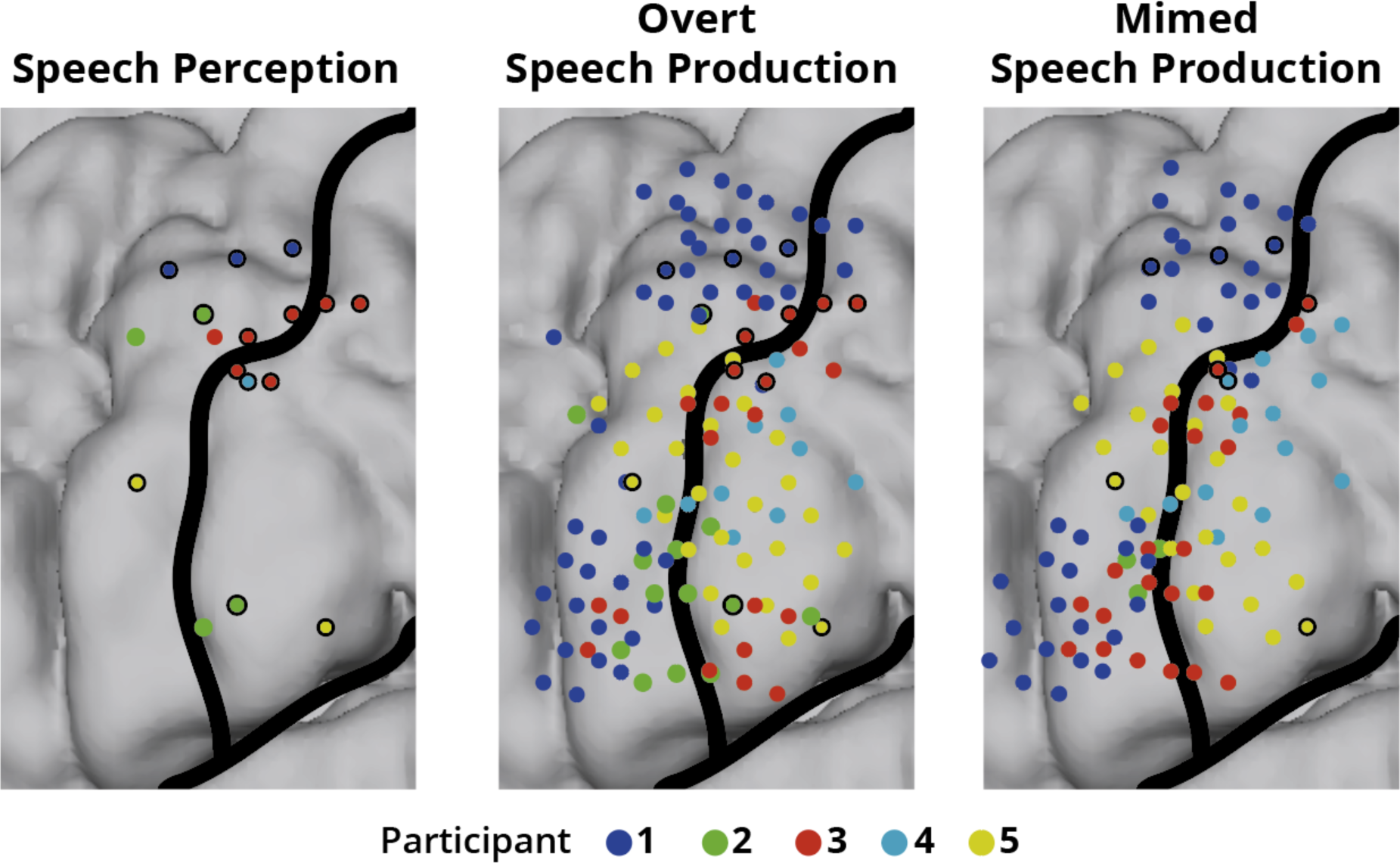
Significantly responding electrodes to perceived (left), overt produced (middle), mimed produced (right) speech over all subjects. Electrodes are visualized on an MNI brain. Central sulcus and sylvian fissure are indicated with black lines. Different participants are indicated in different colors. Speech perception electrodes that are responsive during at least one version of speech production have a black border (left panel), as have speech production electrodes that are also responsive during speech perception (middle and right panel).

### Classification of overt produced speech sounds

SVM was able to accurately classify the overt produced speech sounds for all participants above chance (p < .01, mean chance level 63.42%), with an average decoding accuracy of 85.11% (ranging between 81.47% – 91.07%, Figure 3A). On average 0.72% of the production-rest epochs were classified as a produced sound (production false positive, Figure 3B). Standardizing the epochs based on the mean and standard deviation of the rest periods during the production task resulted in slightly different (but significant above chance, p < .01, mean chance level 53.59%) decoding accuracies: 84.24% (range between 81.03% – 88.93%, an average decrease of 0.87 percentage points, Figure 3C). An average of 0.72% of production-rest epochs were classified as a produced sound after rest normalization (Figure 3D).

**Figure 3:**
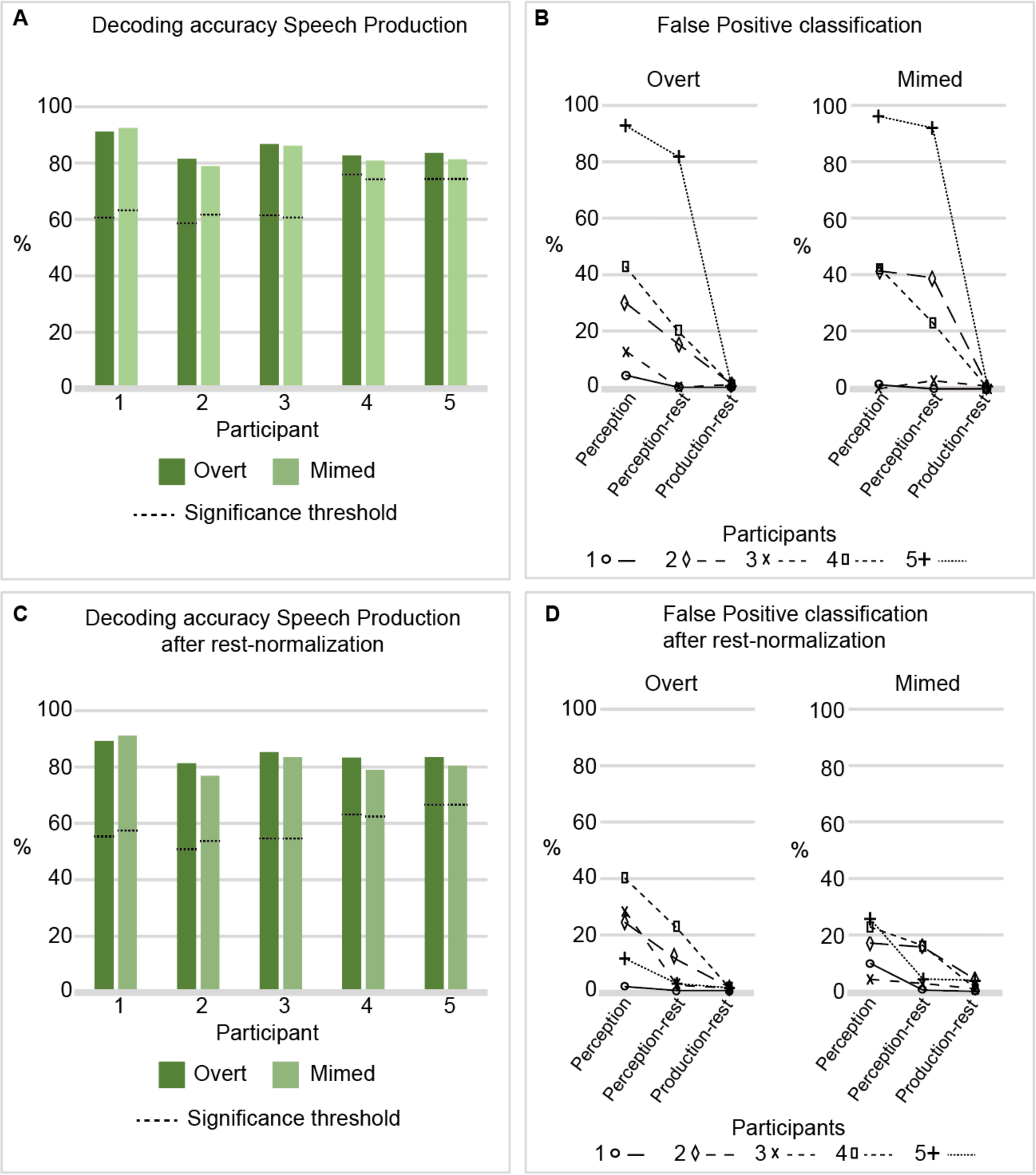
Classification scores and False Positive rates. A, C) Classification scores on self-produced speech epochs for each participant using an overt-based (dark green) and mimed-based (light green) decoder, before (A) and after (C) rest normalization. Dashed lines indicate the significance threshold (p = .05) of the classification accuracy based on the distribution accuracies generated using random labels. B, D) Percentages of false positively classified epochs for the decoder based on the overt and mimed speech separately, before (B) and after (D) rest normalization. The percentage scores of false positive classified epochs are visualized for the perception epochs (left), the perception-rest epochs (middle), and the production-rest epochs (right). Subjects are indicated with different colors and markers.

### Classification of mimed produced speech sounds

The mimed produced sounds were classified with an average classification accuracy of 83.90% (range 78.85% – 92.36%, Figure 3A), which was significantly above chance for all participants (p < .01, mean chance level 64.08%). On average 0.82% of the production-rest epochs were classified as false positives (Figure 3B). After rest-normalization, classification accuracy of the mimed sounds was on average 81.93 % (range 76.65% – 90.91%, average decrease of 1.96 percentage points, Figure 3C), which was again above chance (p < .01, mean chance level 54.40%). On average, 2.01% of production-rest epochs were classified as a production false positive (Figure 3D).

### False positive classification during speech perception using a decoder based on overt speech

Applying the overt-based classifier on perception data generated false positives in all participants. On average, 36.57% (range 4.29% - 92.86%) of speech perception epochs were classified as false positives, while an average of 23.34% (range 0.00% – 81.77%) of perception-rest epochs were classified as false positives (Figure 3B). A one-sided Wilcoxon signed rank test confirmed there were significantly more false positives during perception epochs compared to perception-rest epochs (p < .05), but not significantly more during perception-rest compared to production-rest (p = .13). After rest normalization, speech perception epochs were false positively classified in on average 21.14% of epochs (range 1.43% - 40.00%), which was again significantly higher (p < .05) than the false positive classifications during perception rest epochs: 7.50% (range 0.00% - 22.17%, Figure 3D). There were not significantly more false positives during perception-rest compared to production-rest (p = .06). Looking at the consistency across participants, the percentage scores of false positive classified perception epochs decreased after rest normalization in four participants (1, 2, 4, and 5), while it increased for participant 3. The percentage scores of false positive perception-rest epochs remained similar before and after normalization in participant 1, decreased in participants 2 and 5, but increased in participants 3 and 4.

### False positive classification during speech perception using a decoder based on mimed speech

For mimed speech, an average of 36.29% (range 0.00% - 95.71%) of speech perception epochs were classified as false positives, versus 31.34% (range 0.00% - 91.63%) of the perception-rest epochs (Figure 3B). A one-sided Wilcoxon signed rank test found the difference between false positives during perception and perception-rest epochs non-significant (p = .16), nor was the difference between false positives during perception-rest and production-rest (p = .06). After rest normalization, the average percentages false positive classified epochs were 16.00% (range 4.27% - 25.71%) and 7.95% (0.49% - 16.26%) for perception and perception-rest epochs, respectively (Figure 3D). Here, there were significantly more false positives during the perception epochs than during the perception-rest epochs (p < .05), and significantly more false positives during perception-rest than production-rest (p < .05). Rest normalization caused (compared to the scores before normalization) a decrease in percentage score of false positive classified perception epochs in three participants (2, 4, and 5), while these percentage scores increased in the other two (1 and 3). For the perception-rest epochs, the scores remained similar in one participant (3), decreased in three (2,4, and 5), and increased in participant 1.

### Distinguishing speech perception from speech production and rest

A separate SVM was trained and tested on all speech perception, speech production, and rest epochs jointly, by classifying each epoch as a perception, production, or rest epoch using a LoO approach (after rest-normalizing every epoch). The three states were highly separable, with average classification accuracies of 91.68% (range 88.31% - 95.30%) and 90.69% (range 87.34% - 95.07%) for overt and mimed speech, respectively.

## Discussion

Recent developments in speech decoding are very promising for individuals who can no longer speak (Angrick et al., 2023; Luo et al., 2023; Metzger et al., 2022, 2023; Moses et al., 2021; Wairagkar et al., 2023; Willett et al., 2023). However, those results are typically achieved in controlled lab settings, and not in the daily lives of intended end users. Testing speech BCI reliability in real-life settings is the next hurdle the field needs to take to ensure that BCIs can successfully be adopted by paralyzed individuals. Here, we simulated the scenario where a BCI user is perceiving speech (an intrinsic part of communication) and demonstrate that the perception of speech can be a source of false positive activations of a speech BCI that is trained on produced speech.

The current results confirm that the area frequently used as a signal source for speech decoding BCIs, the ventral sensorimotor cortex, also becomes activated during the perception of speech, as was demonstrated before (Cheung et al., 2016; Pulvermüller et al., 2006; Rhone et al., 2016; Skipper et al., 2005; Venezia et al., 2021). While sensorimotor activation during speech perception is not as widespread as it is during speech production, the spatial activity patterns during perception resemble those during produced speech enough for a speech decoder to identify perceived words as produced words. This was true for decoders based on both overt and mimed speech. The results show that these false positives arise more during speech perception than during rest periods in the perception task (except when using the mime-decoder without doing rest normalization).

Calibration of the brain signals is an often-used practice before conducting a BCI session and aims to normalize the incoming brain signals based on brain signals acquired before a BCI experiment. In the current study, calibration using a three-minute rest task recorded independently from the speech tasks did not prevent the occurrence of considerable numbers of false positive classifications due to perceived speech. The argument can be made that these false positives are the result of day-to-day variations in the brain signals (as the speech-production and speech-perception tasks were not necessarily acquired on the same day), which could explain the higher number of false positives during perception-rest epochs compared to production-rest epochs (though this difference was not significant before rest normalization). However, after accounting for the day-to-day variations by normalizing the data on the task-specific rest intervals, the issue of false positive classifications was not resolved. The current results show that this approach to calibration has different effects on the data of different individuals, where for some participants the occurrence of false positives even increased.

To be able to limit the occurrence of false positives caused by speech perception, speech decoders need to be able to distinguish signals corresponding with speech perception from those during production. In the current study there were considerable differences in activity patterns between the speech perception and speech production data. First, activity during speech production was relatively widespread over the SMC, while during perception fewer SMC sites were engaged. Second, perception and production could be distinguished from each other and from rest quite well, as demonstrated by a three-way SVM, with up to 95.30% correct classification. This suggests that the occurrence of false positives during speech perception could potentially be reduced, or even completely removed, if activity patterns during speech perception are considered in the design of speech decoders.

One potential way to decrease the occurrence of perception false positives would be to implement a three-step approach, where after the neural detection of speech, the brain activity patterns are classified as being self-produced or perceived, and only those classified as self-produced are transferred to the decoder that classifies the actual speech content. Another strategy may be to train a decoder that includes speech perception as an additional class, such that neural signal patterns associated with speech perception may be decoded as this class (which could be extended for other sources of false positives). Both strategies would require the acquisition of additional data for decoder design and training or may need to be updated on a regular basis or extended with additional sources of false positives. Whichever strategy is chosen, the additional burden will be worthy in the long run if this ensures a more reliable speech BCI in a daily-life setting.

The current study clearly illuminates speech perception as a source of false positive activation of a speech BCI, but there are some limitations to this work. First of all, the number of trials on which the decoders are trained are rather small. More trials may be beneficial in decreasing the false positive rates, but this requires further testing. Furthermore, this study is limited to high-frequency signals, which has proven to be a reliable source of speech-related activation, but speech decoding may benefit from a combination of frequency bands (see for example Metzger et al., 2022). The question remains how the occurrence of false positives is impacted by including signal from different frequency bands. A third limitation is the fact that we classified single syllables, rather than full words. Longer words benefit from variable word lengths which may make them easier distinguishable from neural data (or harder to detect in case of speech perception). However, the current findings may be especially relevant for speech BCIs that allow for spelling based on single phones or phonemes. Fourth, we are classifying segments of neural data based on behavioral events (sound or movement onset), rather than detecting speech events based on the neural data itself. Further research is necessary with speech detection algorithms based on neural events to determine whether the occurrence of false positives during speech perception is similar in such situations. Finally, perhaps the most important limitation is that this work is done with abled individuals. It is of utmost importance to investigate how these findings translate to paralyzed users.

The likelihood of a speech BCI being adopted into daily-life depends, among others, on the experienced reliability of the device (Kübler et al., 2014). If speech BCIs are designed with real-life situations in mind and show high reliability also during two-way conversation, they have a higher likelihood of being embraced by end-users and thus provide added value. With this work, we hope to stimulate those who are developing (speech) BCIs to test their systems in naturalistic settings and incorporate steps to decrease the occurrence of false positives generated by external sources.

## Competing interests

The authors declare no competing interests.

## Funding statement

This study is supported by the Dutch Technology Foundation STW (grant UGT7685), Dutch Science Foundation (SGW-406-18-GO-086), European Research Council (ERC-Advanced ‘iConnect’ project, grant ADV 320708), and the National Institute on Deafness and Other Communication Disorders (U01DC016686) and National Institute of Neurological Disorders and Stroke (UH3NS114439) of the National Institutes of Health.

## Data availability statement

Data is not made publicly available because not all participants consented with public sharing of their (anonymous) data. Reasonable requests for sharing of the data can be made to the corresponding author.

## Ethics statement

The Medical Ethical Committee of the University Medical Center Utrecht gave ethical approval for this work.

## Notes

### Competing Interest Statement

The authors have declared no competing interest.

